# BIRTH SPACING AND UNDER-FIVE CHILD SURVIVAL IN GHANA: EVIDENCE FROM THE 2022 DEMOGRAPHIC AND HEALTH SURVEY

**DOI:** 10.64898/2026.10.14.26365524

**Authors:** Abubakar Iddrisu Siddiq, Juliet Akpene Akuffo, Adu Gyamfi Oppong, Anne-Marie Xoese Ditchfield, Sarah Agyarko Duah, Gideon Lantei Lamptey

## Abstract

**Background:** Short birth intervals are a well-documented, modifiable risk factor for child death in low- and middle-income countries, yet nationally representative evidence from Ghana’s most recent Demographic and Health Survey has not been fully examined. This study assessed the association between the preceding birth interval and under-five mortality using the 2022 Ghana Demographic and Health Survey.

**Methods:** The analysis used the complete birth history reported by women aged 15-49 years. The cohort comprised births occurring 60-119 completed months (5-10 years) before the survey, which allowed a complete five-year observation window for under-five survival (n = 8,418; 397 deaths). Spacing models were restricted to second-or-higher-order births with a defined preceding interval (n = 6,340; 282 deaths), categorised as under 18, 18-23, 24-35 (reference), 36-47, and 48 or more months. Crude and adjusted odds ratios were estimated using survey-weighted, Taylor-linearised logistic regression, adjusting for birth order, child sex, plurality, maternal age, residence, region, maternal education, wealth quintile, and marital status.

**Results:** Weighted under-five mortality in the cohort was 45.4 per 1,000 live births (4.54%). Among second-or-higher-order births, weighted mortality fell from 9.3% after intervals of under 18 months and 8.0% after 18-23 months to 4.2%, 3.2%, and 3.5% after 24-35, 36-47, and 48 or more months, respectively. Relative to 24-35 months, the adjusted odds of death were higher after intervals of under 18 months (adjusted odds ratio [AOR] 2.58, 95% confidence interval [CI] 1.48-4.49, p < 0.001) and 18-23 months (AOR 2.09, 95% CI 1.33-3.29, p = 0.001); longer intervals did not differ from the reference. Multiple birth (AOR 2.48, 95% CI 1.33-4.63) and having a mother who was never in union (AOR 3.25, 95% CI 1.29-8.17) were also associated with higher odds of death, and marked regional variation persisted after adjustment.

**Conclusion:** A short preceding birth interval, particularly one of under 18 months, was independently associated with an approximately two-to two-and-a-half-fold increase in the odds of under-five death in this nationally representative sample. The findings support continued investment in postpartum family planning to help women space births at least 24 months apart, alongside closer attention to multiple births, unmarried mothers, and the regions carrying the greatest burden.

## 1. Introduction

Under-five mortality remains one of the clearest markers of a country’s health and development. Ghana has recorded a long-term decline in child deaths over the past two decades, yet the rate reported in the 2022 Ghana Demographic and Health Survey still leaves the country short of the Sustainable Development Goal target of 25 or fewer deaths per 1,000 live births by 2030 (1). Children in this age group are the most vulnerable to infection, undernutrition, and the consequences of inadequate maternal and newborn care. Identifying determinants that are both strongly associated with survival and realistically modifiable through programme action is therefore a continuing priority for Ghanaian public health.

The interval between successive live births is one such determinant. A child born soon after an older sibling enters a household in which the mother’s physiological reserves, including her iron stores and nutritional status, may not yet have recovered from the preceding pregnancy and lactation — a pattern often described as maternal depletion. A short interval also compresses the period during which breastfeeding, a strong protective factor against infant infection, can continue before the next pregnancy displaces it, and it increases direct competition between closely spaced siblings for a household’s food, care, and attention. Younger siblings born into a household that already contains another very young child are additionally exposed to a higher risk of person-to-person transmission of common childhood infections. These mechanisms have been described in detail in earlier analyses of Demographic and Health Survey data from dozens of countries (2,3), and have since been corroborated in meta-analyses linking short intervals to low birthweight, preterm birth, and elevated neonatal and infant mortality (4,5).

In 2005, the World Health Organization convened a technical consultation that recommended an interval of at least 24 months between a live birth and the next pregnancy — equivalent to a birth-to-birth interval of roughly 33 months — largely on the strength of this mortality and morbidity evidence (6,7). That recommendation was later reinforced in global guidance on postpartum family planning programming (8,9). Subsequent pooled analyses using Demographic and Health Survey data from across sub-Saharan Africa, Asia, and Latin America have found the association between short intervals and child death to be broadly consistent across settings, although its magnitude varies by country and by the length of interval examined (10). An eight-country West African analysis that included Ghana reported a similarly elevated risk of under-five death after short preceding intervals (11), and a recent study in rural Tanzania reported comparable effects using detailed demographic surveillance data (12).

Within Ghana specifically, an analysis of household survey data on maternal socioeconomic predictors found that short birth intervals disproportionately affected women with less education and lower household wealth and were associated with poorer child survival outcomes (13). Ghana’s under-five mortality has also been shown to vary considerably by wealth, maternal education, region, and residence (14), raising the possibility that any observed association between birth spacing and child death could partly reflect these wider social and geographic inequalities rather than birth spacing itself. A properly specified multivariable analysis that adjusts for these factors is therefore needed before the independent contribution of birth spacing can be stated with confidence.

The 2022 Ghana Demographic and Health Survey, fielded under national authorisation and analysed here under a project registered with the DHS Program (authorisation granted 3 September 2026), offers the most recent nationally representative data with which to revisit this question. Its complete birth history, covering every live birth reported by women aged 15–49 years, makes it possible to reconstruct a cohort of children who have already passed through the age window in which under-five death can occur, while recording the length of the interval that preceded each birth. To date, published analyses of the 2022 round have examined the predictors and timing of under-five death more generally (15); no published analysis has yet used this round specifically to re-examine the birth spacing and under-five mortality relationship in Ghana with a fully survey-weighted multivariable model.

This study therefore had two objectives. The first was to describe the distribution of the preceding birth interval and of under-five mortality among births occurring 5–10 years before the 2022 Ghana Demographic and Health Survey. The second was to estimate the association between the preceding birth interval and the odds of under-five death, before and after adjustment for birth order, child sex, plurality, maternal age, residence, region, maternal education, household wealth, and marital status. In doing so, the study aimed to provide contemporary, nationally representative evidence to inform family planning and child survival programming in Ghana. By providing updated estimates of these risks, this analysis addresses the critical need to identify which demographic and socioeconomic factors continue to drive disparate health outcomes for children in the region (11,16). Furthermore, by clarifying the magnitude of these risks within the current socio-economic landscape, the findings offer a robust evidence base for tailoring interventions to populations where short birth intervals remain highly prevalent (13,17). By employing discrete-time survival analysis with appropriate survey weighting, this investigation seeks to enhance the precision of risk estimates associated with varying birth intervals (18). Given that the prevalence of short birth intervals remains a significant predictor of mortality, identifying these associations is crucial for targeting health interventions effectively (19). Furthermore, by accounting for potential confounding through rigorous statistical adjustment, this research clarifies whether the documented mortality risks persist independently of the underlying socioeconomic inequalities that define the current Ghanaian landscape (20).

## 2. Methods

### 2.1 Study design and setting

This study used the 2022 Ghana Demographic and Health Survey, a nationally representative cross-sectional survey, and analysed it in the form of a retrospective birth cohort. Reporting follows the Strengthening the Reporting of Observational Studies in Epidemiology statement for cross-sectional studies, adapted to the retrospective birth-history design that is standard in Demographic and Health Survey mortality analyses (21).

### 2.2 Data source

Data were drawn from the Births Recode file of the 2022 Ghana Demographic and Health Survey, which was implemented by the Ghana Statistical Service with technical support from the DHS Program (22). The Births Recode file contains one record for every live birth reported in the complete birth history of interviewed women aged 15–49 years, including the date of birth, survival status, age at death if applicable, and the interval since the preceding birth. Access to the dataset for this registered research project was authorised by the DHS Program on 3 September 2026.

### 2.3 Sampling design

The Ghana Demographic and Health Survey uses a stratified, two-stage cluster sampling design. Enumeration areas are selected as primary sampling units within sampling strata formed by crossing region with urban or rural residence, and households are then systematically selected within each enumeration area (23). All eligible women aged 15–49 years in the selected households were interviewed, and each woman’s complete birth history was recorded. As with Demographic and Health Surveys worldwide, this design departs from simple random sampling by introducing clustering, stratification, and unequal selection probabilities, and it therefore requires analytic methods that account for these features, since analyses that ignore them can yield biased point estimates and underestimated standard errors (24,25).

### 2.4 Study population and analytic sample

The Births Recode file contained 34,663 live births reported by interviewed women. For a child’s under-five survival status to be fully and unambiguously observed, the child must already have had the opportunity to reach five years of age by the time of interview; children born more recently have not yet completed this observation window. The analytic cohort was therefore restricted to births occurring 60–119 completed months (5–10 years) before the survey date, yielding 8,418 births, among which 397 under-five deaths were recorded. First-order births, for which no preceding sibling exists and the preceding birth interval is therefore undefined, numbered 2,078. These were retained for the descriptive characterisation of the cohort but necessarily excluded from the birth-spacing regression models, leaving 6,340 second-or-higher-order births (282 deaths) for those models. The flow of births through these inclusion criteria is shown in Figure 1.

**Figure 1.**
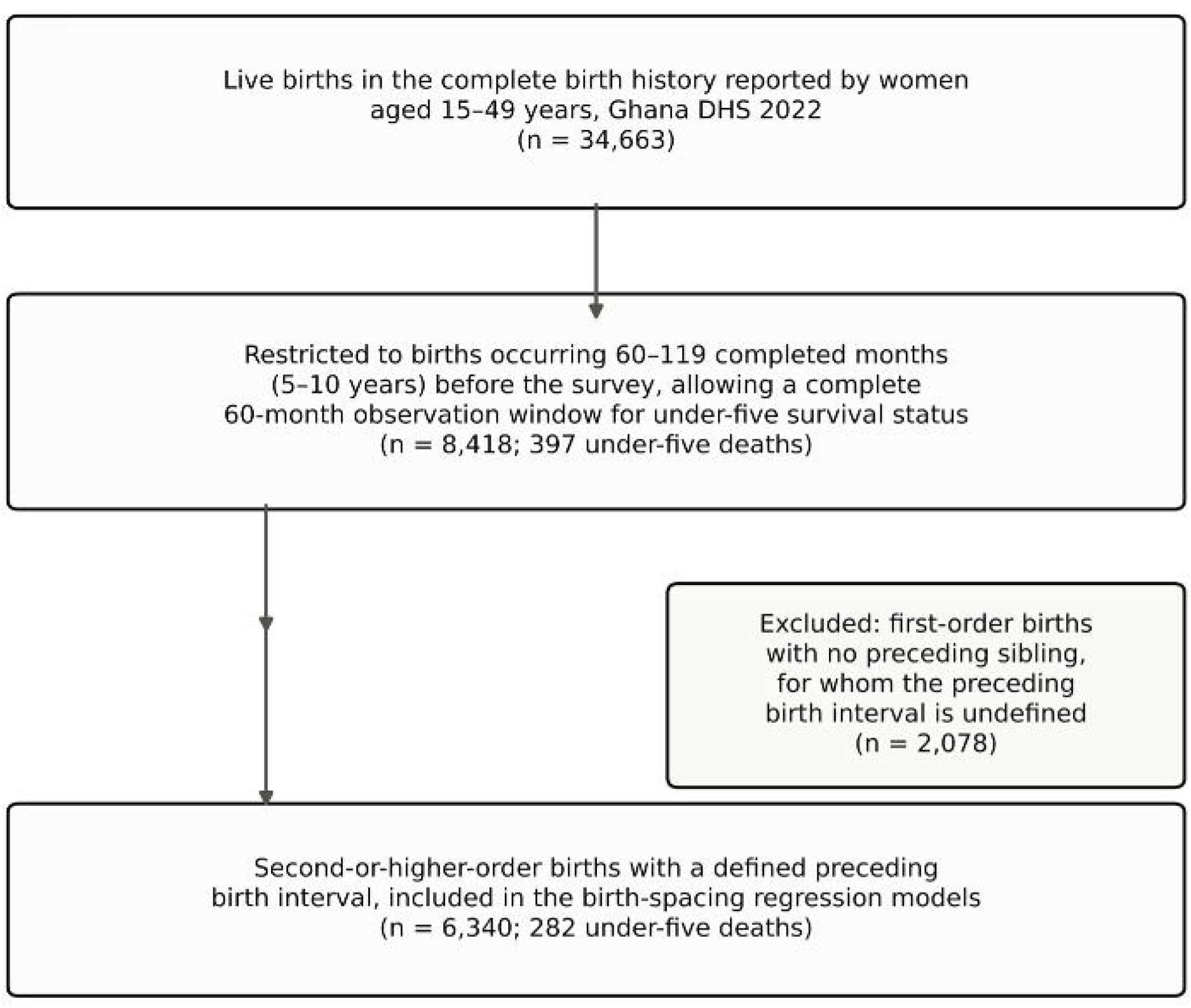
Flow diagram of birth inclusion in the analytic cohort

### 2.5 Outcome variable

The primary outcome was death before the fifth birthday (under-five death), a binary variable derived from the child’s reported survival status and, for children who had died, their age at death in completed months. A child was classified as having died before age five if the survival status indicated death and the age at death was under 60 months. All other children in the analytic cohort, by construction, had already survived to at least 60 months of age. This restriction avoids the immortal-time bias that would arise from applying a five-year mortality definition to children who have not yet lived long enough for the outcome to be observable.

### 2.6 Exposure variable

The primary exposure was the preceding birth interval, defined by the Demographic and Health Survey as the number of completed months between a birth and the birth immediately preceding it to the same mother. Following the World Health Organization’s recommendation of an inter-pregnancy interval of at least 24 months (6) and the categorisation used in comparable analyses (2,10,11), the interval was grouped into five categories: under 18 months, 18–23 months, 24–35 months (the reference category), 36–47 months, and 48 months or more.

### 2.7 Covariates

Covariates were selected on the basis of established associations with child survival in the Ghanaian and wider sub-Saharan African literature. They comprised: birth order, grouped as first, second or third, fourth or fifth, and sixth or higher; sex of child; plurality, distinguishing a single birth from a multiple (twin or higher-order) birth; maternal age at the birth of the child, grouped as under 20, 20–34, and 35 years or older; place of residence (urban or rural); region of residence (16 regions); maternal educational attainment, grouped as no education, primary, secondary, and higher; household wealth quintile, constructed by the DHS Program from household asset data; and maternal marital status at the time of interview, grouped as currently in union, formerly in union, and never in union. Biomedical covariates such as antenatal care attendance, place of delivery, and birthweight were considered but not included, because these variables are recorded only for births in the five years immediately preceding the survey and are therefore largely unavailable for the 5–10-year birth cohort used here. This constraint is addressed further as a limitation.

### 2.8 Statistical analysis: descriptive statistics

Descriptive characteristics of the analytic cohort were summarised as weighted percentages, applying the DHS-constructed sampling weight after dividing it by its documented scaling factor of 1,000,000. Weighted under-five mortality was calculated overall and separately within each level of every covariate, using the same sampling weight.

### 2.9 Statistical analysis: bivariate associations

Bivariate (crude) associations between each covariate, including the preceding birth interval, and under-five death were estimated using separate survey-weighted logistic regression models, each containing a single categorical predictor. Models were fitted using Taylor-linearised variance estimation that incorporated the primary sampling unit, sampling stratum, and sampling weight, producing crude odds ratios with 95% confidence intervals and Wald test p-values. Specifying the sampling units, strata, and weights in this way allows the variance estimates to reflect the complex design rather than assuming simple random sampling (26).

### 2.10 Statistical analysis: multivariable regression and model diagnostics

A multivariable survey-weighted logistic regression model was fitted with under-five death as the outcome and all covariates listed above entered simultaneously alongside the preceding birth interval, again using Taylor-linearised variance estimation that accounted for the survey’s stratification and clustering. Adjusted odds ratios, 95% confidence intervals, and p-values were obtained from this model. Multicollinearity among covariates was assessed using the variance inflation factor, calculated on an unweighted, fully indicator-coded design matrix. All values were below the conventional threshold of 10, with the highest observed for the no-education category of maternal education (variance inflation factor 8.98), indicating that collinearity did not materially distort the adjusted estimates. All analyses were conducted in Python, using the samplics package for survey-weighted generalised linear models and the statsmodels package for the variance inflation factor diagnostic.

### 2.11 Handling of missing data

Births with a missing value for any covariate included in a given model, or for the sampling design variables required for weighting, were excluded from that model through listwise deletion; the number of births contributing to each model is reported with its corresponding results. First-order births were excluded from the birth-spacing regression models by design, as described above, rather than through missingness, since the preceding birth interval does not exist for a first birth rather than being unrecorded.

### 2.12 Ethical considerations

The 2022 Ghana Demographic and Health Survey received ethical clearance from the Ghana Health Service Ethics Review Committee and from the Institutional Review Board of ICF in the United States, and written informed consent was obtained from all respondents at the time of data collection. The data used in this study are fully de-identified, contain no household addresses or names, and were accessed for this registered secondary analysis under an authorisation granted by the DHS Program on 3 September 2026, following the DHS Program’s standard data-use terms.

## 3. Results

### 3.1 Characteristics of the analytic sample

Of the 34,663 live births recorded in the complete birth history, 8,418 occurred 60–119 completed months before the survey and formed the analytic cohort, of which 397 ended in death before age five. Among these births, 24.8% were first-order births, 40.0% were second or third order, 23.3% were fourth or fifth order, and 11.9% were sixth order or higher. Slightly more than half of the children were male (50.8%), and 3.9% were part of a multiple birth. Most mothers (72.6%) were aged 20–34 years at the birth of the child, 50.9% of births occurred in rural areas, and Ashanti (18.6%) and Greater Accra (12.9%) contributed the largest weighted shares of births. Close to three in ten mothers (29.7%) had no formal education, and household wealth was fairly evenly distributed across the five quintiles, from 18.1% in the richest to 22.9% in the poorest. Among the 6,340 second-or-higher-order births with a defined preceding interval, 5.5% followed an interval of under 18 months, 9.7% an interval of 18–23 months, 29.3% an interval of 24–35 months, 21.4% an interval of 36–47 months, and 34.2% an interval of 48 months or more.

### 3.2 Under-five mortality by birth interval and other characteristics

The overall weighted under-five mortality rate in the analytic cohort was 45.4 deaths per 1,000 live births (4.54%). Weighted under-five mortality varied sharply across categories of the preceding birth interval, from 9.3% after an interval of under 18 months and 8.0% after 18–23 months, to 4.2% after 24–35 months, 3.2% after 36–47 months, and 3.5% after 48 months or more (Table 1; Figure 2). Risk therefore declined steeply as the interval lengthened from under 18 months to 36–47 months, with no further decline at intervals of 48 months or more. Weighted under-five mortality was also higher among multiple births (10.4%, compared with 4.3% for singletons), among births to mothers aged under 20 years (7.2%), among births of sixth order or higher (6.0%), and among births to mothers who had never been in a union (6.9%). Mortality varied considerably by region, from 1.8% in Greater Accra to 7.7% in Volta region and 7.6% in Northern region, and was higher in the poorest wealth quintile (6.0%) than in the richest (3.0%).

**Figure 2.**
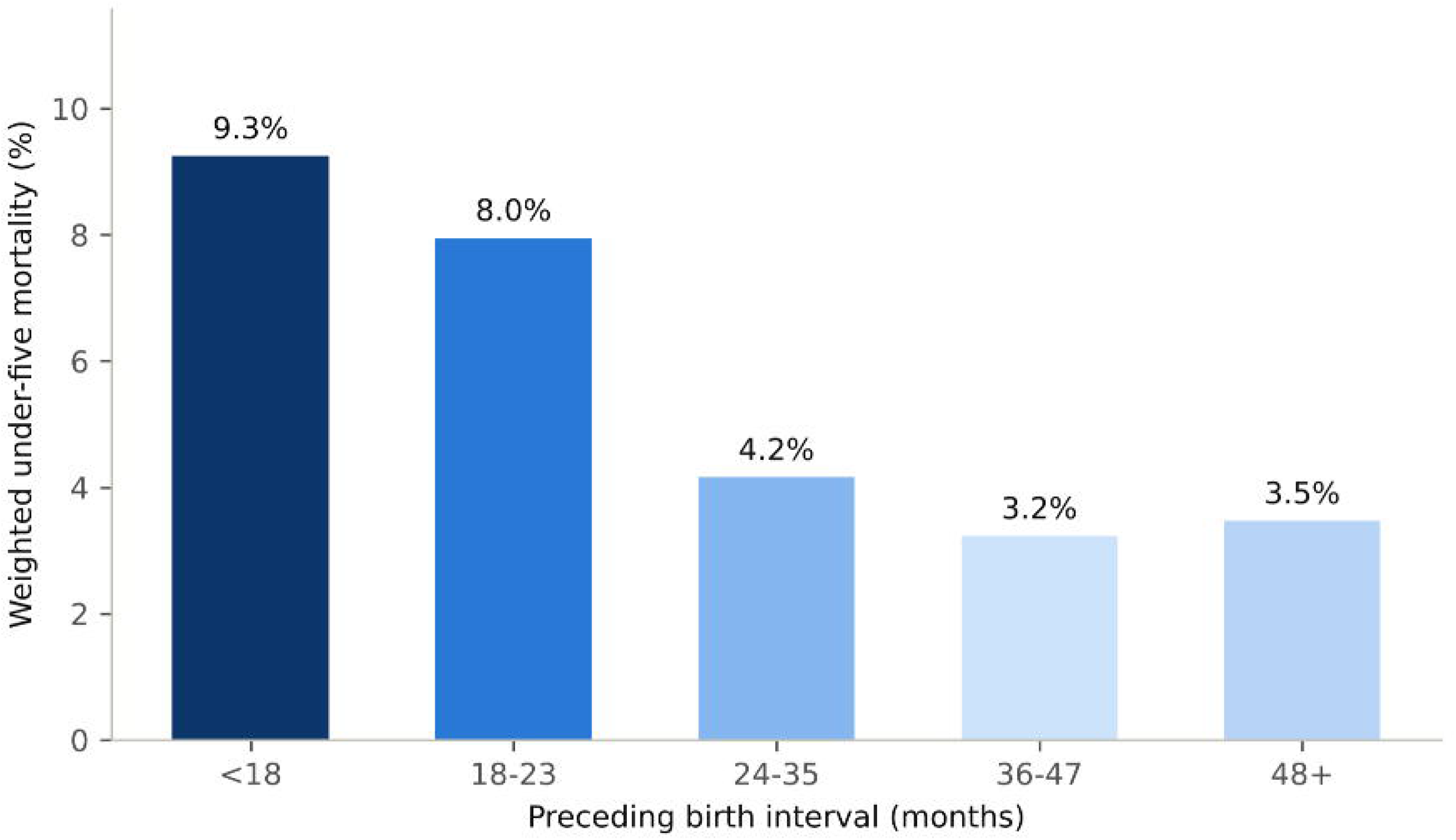
Under-five mortality by preceding birth interval among births 5–10 years before the survey, Ghana DHS 2022

**Table 1.** Weighted characteristics of the analytic cohort (births 5–10 years before the survey, n = 8,418) and weighted under-five mortality by characteristic, Ghana DHS 2022.

| Characteristic | Level | n | Weighted | Under-5 mortality, |
| --- | --- | --- | --- | --- |

|  |  | (unweighted) | % | weighted % |
| --- | --- | --- | --- | --- |
| Preceding birth interval<br>(months) | <18 | 320 | 5.5 | 9.3 |
|  | 18–23 | 622 | 9.7 | 8.0 |
|  | 24–35 | 1,924 | 29.3 | 4.2 |
|  | 36–47 | 1,428 | 21.4 | 3.2 |
|  | 48+ | 2,046 | 34.2 | 3.5 |
| Birth order | 1 | 2,048 | 24.8 | 4.8 |
|  | 2–3 | 3,249 | 40.0 | 4.0 |
|  | 4–5 | 2,037 | 23.3 | 4.4 |
|  | 6+ | 1,084 | 11.9 | 6.0 |
| Sex of child | Female | 4,131 | 49.2 | 4.0 |
|  | Male | 4,287 | 50.8 | 5.1 |
| Birth type | Multiple birth | 350 | 3.9 | 10.4 |
|  | Single birth | 8,068 | 96.1 | 4.3 |
| Mother's age at birth of<br>child (years) | <20 | 904 | 10.5 | 7.2 |
|  | 20–34 | 6,124 | 72.6 | 4.1 |
|  | 35+ | 1,390 | 16.9 | 4.6 |
| Place of residence | Rural | 4,918 | 50.9 | 4.9 |
|  | Urban | 3,500 | 49.1 | 4.2 |
| Region | Ahafo | 402 | 3.4 | 3.7 |
|  | Ashanti | 567 | 18.6 | 5.0 |
|  | Bono | 427 | 2.9 | 3.9 |
|  | Bono East | 464 | 2.2 | 2.9 |
|  | Central | 475 | 10.2 | 4.0 |
|  | Eastern | 400 | 7.2 | 4.0 |
|  | Greater Accra | 436 | 12.9 | 1.8 |
|  | North East | 695 | 2.9 | 6.2 |
|  | Northern | 586 | 3.4 | 7.6 |
|  | Oti | 513 | 2.7 | 3.2 |
|  | Savannah | 894 | 11.1 | 5.1 |
|  | Upper East | 691 | 2.8 | 4.0 |
|  | Upper West | 484 | 4.1 | 6.8 |
|  | Volta | 410 | 4.4 | 7.7 |
|  | Western | 414 | 6.3 | 5.0 |
|  | Western North | 560 | 4.8 | 5.1 |
| Mother's highest education | No education | 3,275 | 29.7 | 5.3 |
|  | Primary | 1,454 | 16.5 | 5.0 |
|  | Secondary | 3,184 | 46.6 | 4.4 |
|  | Higher | 505 | 7.3 | 1.4 |
| Household wealth quintile | Poorest | 2,721 | 22.9 | 6.0 |
|  | Poorer | 2,042 | 20.7 | 4.4 |
|  | Middle | 1,507 | 19.7 | 5.3 |
|  | Richer | 1,182 | 18.7 | 3.5 |
|  | Richest | 966 | 18.1 | 3.0 |
| Mother's marital status | Currently in union | 7,331 | 84.2 | 4.4 |
|  | Formerly in union | 765 | 11.1 | 4.6 |
|  | Never in union | 322 | 4.7 | 6.9 |

*Note: Preceding birth interval rows are restricted to the 6,340 second-or-higher-order births with a defined interval; all other rows describe the full analytic cohort of 8,418 births. Percentages are weighted and may not sum to 100 because of rounding.*

### 3.3 Bivariate associations with under-five death

In bivariate survey-weighted logistic regression, the odds of under-five death were higher after a preceding interval of under 18 months (crude odds ratio [COR] 2.35, 95% CI 1.36– 4.03, p = 0.002) and after an interval of 18–23 months (COR 1.99, 95% CI 1.26–3.14, p = 0.003), relative to the 24–35-month reference category; intervals of 36–47 months (COR 0.77, 95% CI 0.50–1.20, p = 0.25) and 48 months or more (COR 0.83, 95% CI 0.53–1.29, p = 0.41) did not differ significantly from the reference. A multiple birth (COR 2.41, 95% CI 1.37–4.22, p = 0.002), birth order of six or higher (COR 1.59, 95% CI 1.11–2.28, p = 0.012), and a mother who had never been in a union (COR 2.78, 95% CI 1.23–6.27, p = 0.014) were each associated with higher crude odds of death, as was residence in several regions relative to Greater Accra, most markedly Volta region (COR 5.06, 95% CI 2.03–12.59, p < 0.001) and Northern region (COR 4.85, 95% CI 1.87–12.56, p = 0.001). Higher maternal education and greater household wealth were associated with lower crude odds of death, reaching statistical significance for higher education (COR 0.25, 95% CI 0.08–0.81, p = 0.021) and the richest wealth quintile (COR 0.44, 95% CI 0.21–0.91, p = 0.026) (full results in Table 2).

**Table 2.**
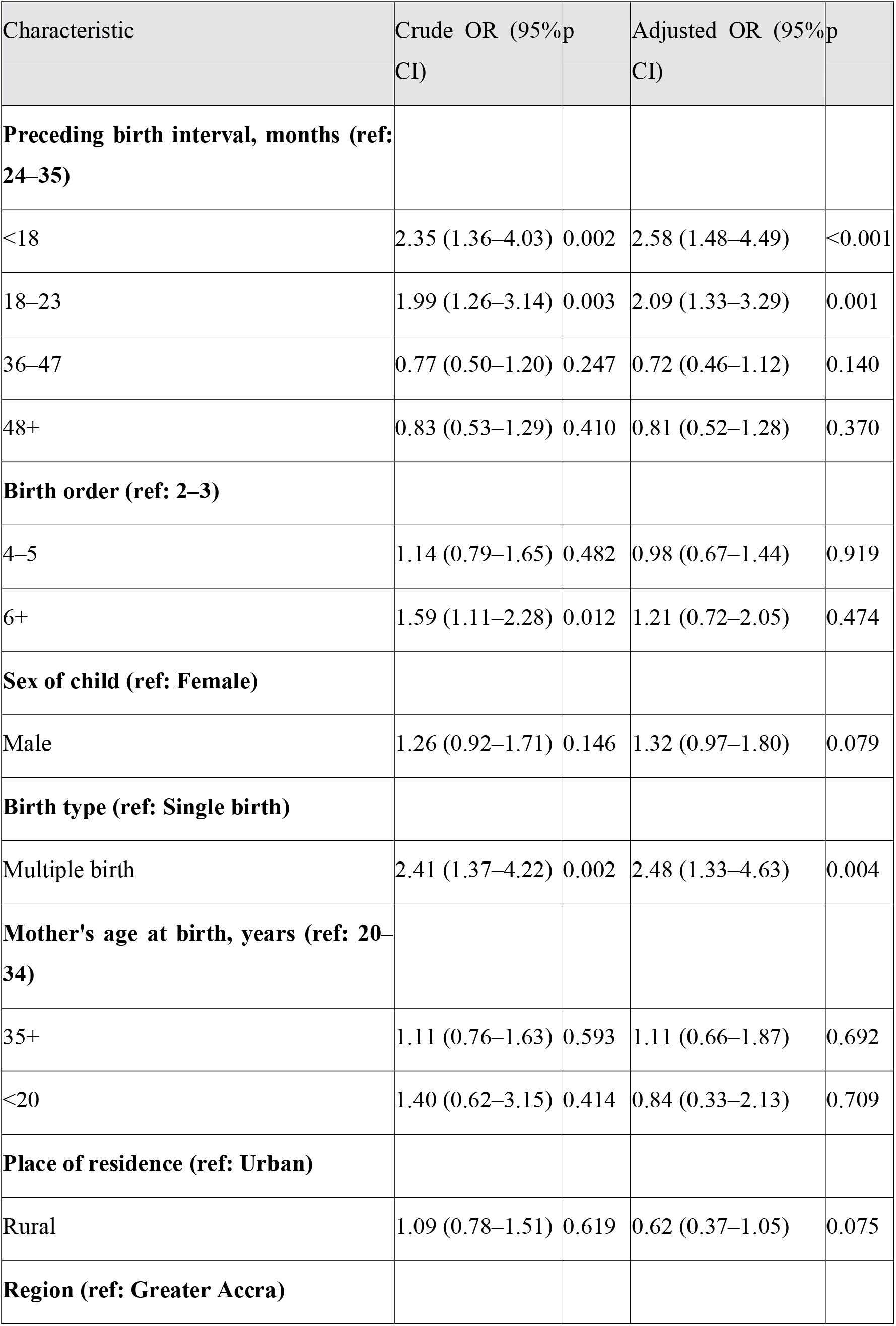
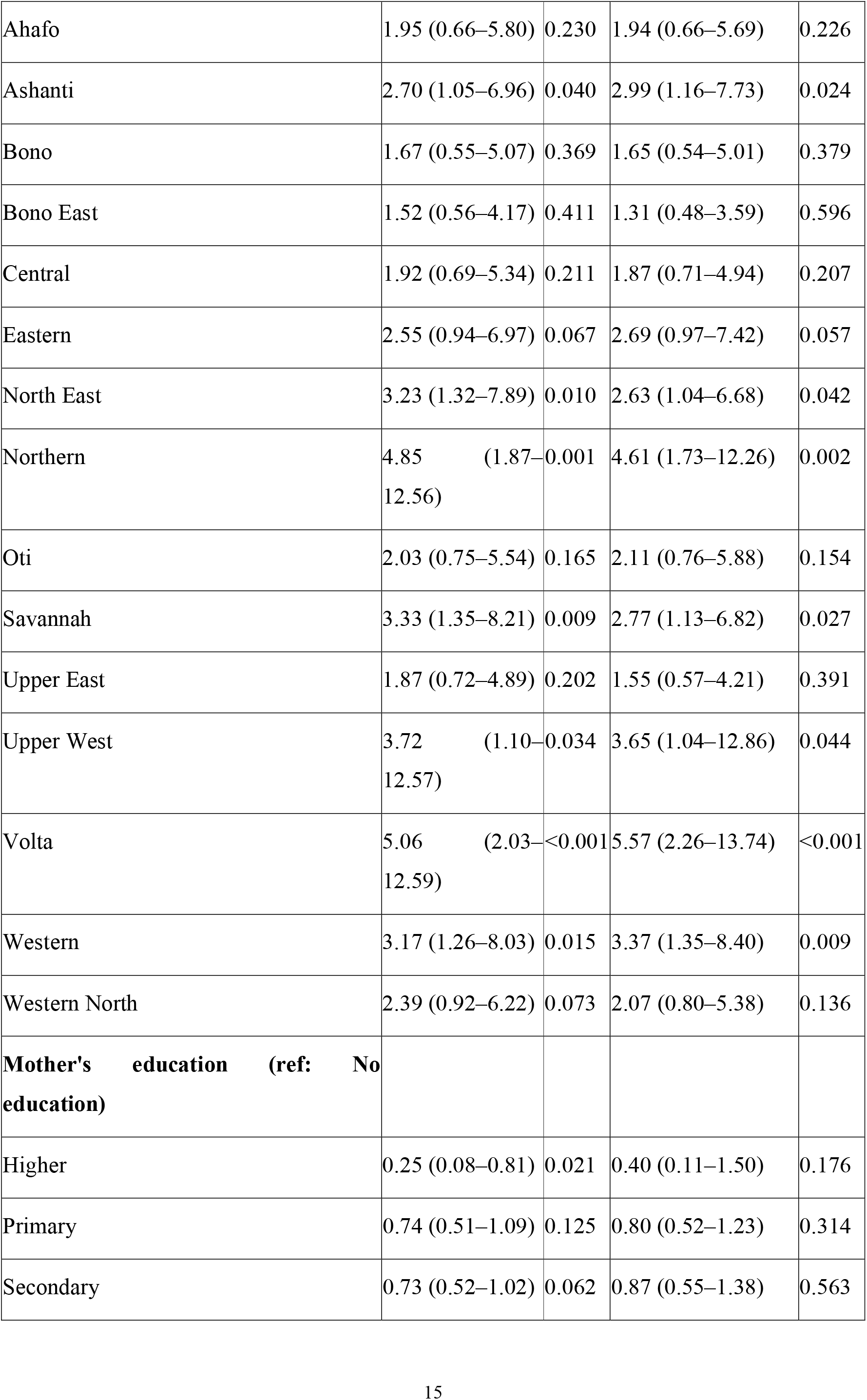
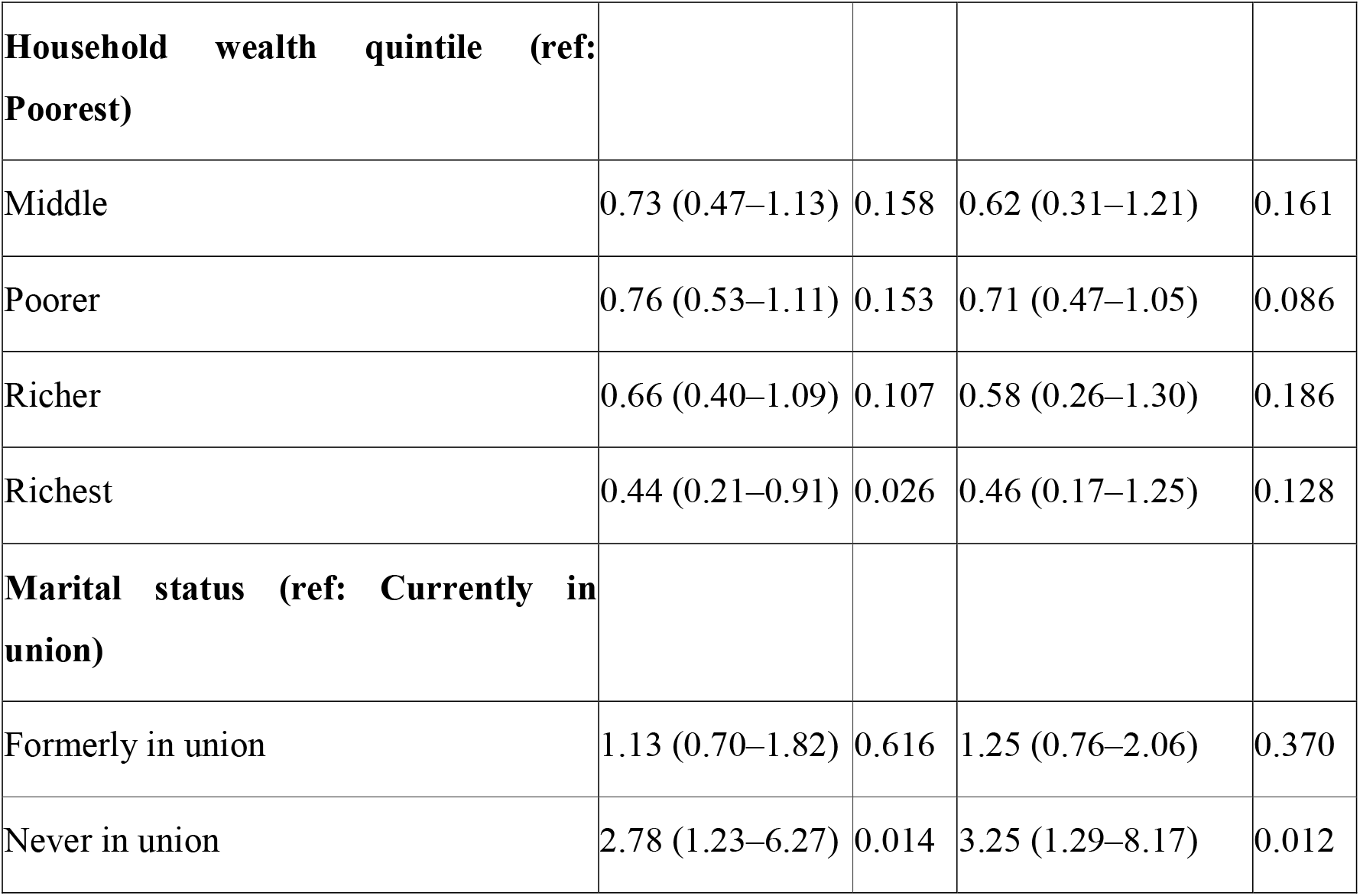
Crude and adjusted odds ratios for under-five death, births with a defined preceding birth interval (n = 6,340; 282 deaths), Ghana DHS 2022.

| Characteristic | Crude OR (95%CI) | p | Adjusted OR (95%CI) | p |
| --- | --- | --- | --- | --- |
| **Preceding birth interval, months (ref: 24–35)** |  |  |  |  |
| <18 | 2.35 (1.36–4.03) | 0.002 | 2.58 (1.48–4.49) | <0.001 |
| 18–23 | 1.99 (1.26–3.14) | 0.003 | 2.09 (1.33–3.29) | 0.001 |
| 36–47 | 0.77 (0.50–1.20) | 0.247 | 0.72 (0.46–1.12) | 0.140 |
| 48+ | 0.83 (0.53–1.29) | 0.410 | 0.81 (0.52–1.28) | 0.370 |
| **Birth order (ref: 2–3)** |  |  |  |  |
| 4–5 | 1.14 (0.79–1.65) | 0.482 | 0.98 (0.67–1.44) | 0.919 |
| 6+ | 1.59 (1.11–2.28) | 0.012 | 1.21 (0.72–2.05) | 0.474 |
| **Sex of child (ref: Female)** |  |  |  |  |
| Male | 1.26 (0.92–1.71) | 0.146 | 1.32 (0.97–1.80) | 0.079 |
| **Birth type (ref: Single birth)** |  |  |  |  |
| Multiple birth | 2.41 (1.37–4.22) | 0.002 | 2.48 (1.33–4.63) | 0.004 |
| **Mother's age at birth, years (ref: 20–34)** |  |  |  |  |
| 35+ | 1.11 (0.76–1.63) | 0.593 | 1.11 (0.66–1.87) | 0.692 |
| <20 | 1.40 (0.62–3.15) | 0.414 | 0.84 (0.33–2.13) | 0.709 |
| **Place of residence (ref: Urban)** |  |  |  |  |
| Rural | 1.09 (0.78–1.51) | 0.619 | 0.62 (0.37–1.05) | 0.075 |
| **Region (ref: Greater Accra)** |  |  |  |  |
| Ahafo | 1.95 (0.66–5.80) | 0.230 | 1.94 (0.66–5.69) | 0.226 |
| Ashanti | 2.70 (1.05–6.96) | 0.040 | 2.99 (1.16–7.73) | 0.024 |
| Bono | 1.67 (0.55–5.07) | 0.369 | 1.65 (0.54–5.01) | 0.379 |
| Bono East | 1.52 (0.56–4.17) | 0.411 | 1.31 (0.48–3.59) | 0.596 |
| Central | 1.92 (0.69–5.34) | 0.211 | 1.87 (0.71–4.94) | 0.207 |
| Eastern | 2.55 (0.94–6.97) | 0.067 | 2.69 (0.97–7.42) | 0.057 |
| North East | 3.23 (1.32–7.89) | 0.010 | 2.63 (1.04–6.68) | 0.042 |
| Northern | 4.85 (1.87–12.56) | 0.001 | 4.61 (1.73–12.26) | 0.002 |
| Oti | 2.03 (0.75–5.54) | 0.165 | 2.11 (0.76–5.88) | 0.154 |
| Savannah | 3.33 (1.35–8.21) | 0.009 | 2.77 (1.13–6.82) | 0.027 |
| Upper East | 1.87 (0.72–4.89) | 0.202 | 1.55 (0.57–4.21) | 0.391 |
| Upper West | 3.72 (1.10–12.57) | 0.034 | 3.65 (1.04–12.86) | 0.044 |
| Volta | 5.06 (2.03–12.59) | <0.001 | 5.57 (2.26–13.74) | <0.001 |
| Western | 3.17 (1.26–8.03) | 0.015 | 3.37 (1.35–8.40) | 0.009 |
| Western North | 2.39 (0.92–6.22) | 0.073 | 2.07 (0.80–5.38) | 0.136 |
| **Mother's education (ref: No education)** |  |  |  |  |
| Higher | 0.25 (0.08–0.81) | 0.021 | 0.40 (0.11–1.50) | 0.176 |
| Primary | 0.74 (0.51–1.09) | 0.125 | 0.80 (0.52–1.23) | 0.314 |
| Secondary | 0.73 (0.52–1.02) | 0.062 | 0.87 (0.55–1.38) | 0.563 |
| **Household wealth quintile (ref: Poorest)** |  |  |  |  |
| Middle | 0.73 (0.47–1.13) | 0.158 | 0.62 (0.31–1.21) | 0.161 |
| Poorer | 0.76 (0.53–1.11) | 0.153 | 0.71 (0.47–1.05) | 0.086 |
| Richer | 0.66 (0.40–1.09) | 0.107 | 0.58 (0.26–1.30) | 0.186 |
| Richest | 0.44 (0.21–0.91) | 0.026 | 0.46 (0.17–1.25) | 0.128 |
| **Marital status (ref: Currently in union)** |  |  |  |  |
| Formerly in union | 1.13 (0.70–1.82) | 0.616 | 1.25 (0.76–2.06) | 0.370 |
| Never in union | 2.78 (1.23–6.27) | 0.014 | 3.25 (1.29–8.17) | 0.012 |

### 3.4 Multivariable associations with under-five death

After adjustment for birth order, sex of child, plurality, maternal age, residence, region, maternal education, wealth quintile, and marital status, the association between a short preceding birth interval and under-five death persisted and, for the shortest interval, strengthened. The adjusted odds of under-five death were more than two and a half times higher after a preceding interval of under 18 months (adjusted odds ratio [AOR] 2.58, 95% CI 1.48–4.49, p < 0.001) and approximately twice as high after 18–23 months (AOR 2.09, 95% CI 1.33–3.29, p = 0.001), relative to the 24–35-month reference category (Table 2; Figure 3). Intervals of 36–47 months (AOR 0.72, 95% CI 0.46–1.12, p = 0.140) and 48 months or more (AOR 0.81, 95% CI 0.52–1.28, p = 0.370) remained statistically indistinguishable from the reference category.

**Figure 3.**
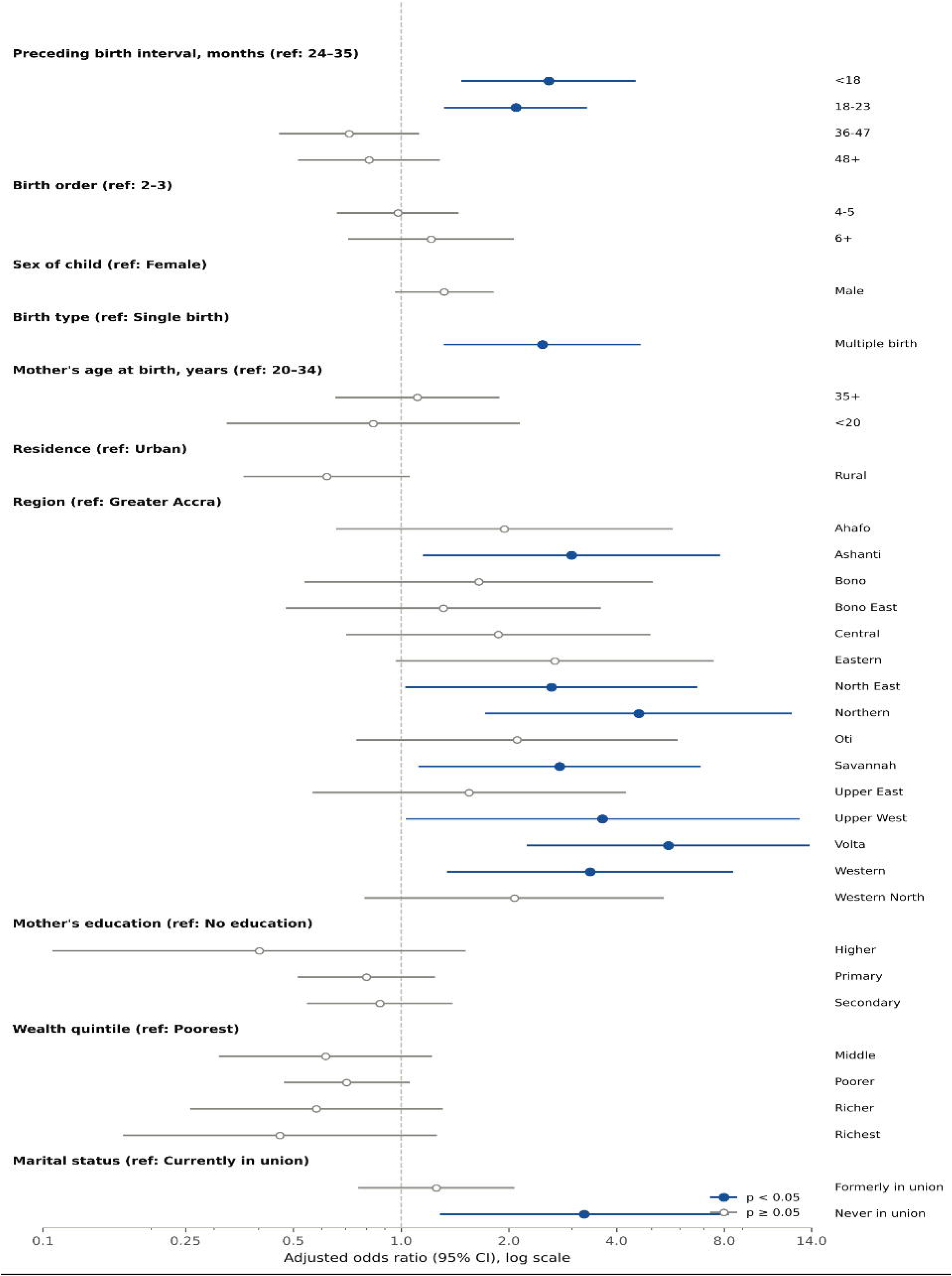
Adjusted odds ratios for birth spacing and other correlates of under-five mortality (multivariable survey-weighted logistic regression, n = 6,340)

A multiple birth remained independently associated with higher odds of death after adjustment (AOR 2.48, 95% CI 1.33–4.63, p = 0.004), as did a mother who had never been in a union (AOR 3.25, 95% CI 1.29–8.17, p = 0.012). Regional variation largely persisted after adjustment: relative to Greater Accra, the adjusted odds of death remained elevated in Volta region (AOR 5.57, 95% CI 2.26–13.74, p < 0.001), Northern region (AOR 4.61, 95% CI 1.73–12.26, p = 0.002), Western region (AOR 3.37, 95% CI 1.35–8.40, p = 0.009), Upper West region (AOR 3.65, 95% CI 1.04–12.86, p = 0.044), Ashanti region (AOR 2.99, 95% CI 1.16–7.73, p = 0.024), Savannah region (AOR 2.77, 95% CI 1.13–6.82, p = 0.027), and North East region (AOR 2.63, 95% CI 1.04–6.68, p = 0.042). Birth order, sex of child, maternal age, rural residence, maternal education, and household wealth quintile were no longer associated with under-five death at conventional levels of statistical significance once the other covariates, including region, were held constant, although point estimates for higher education and greater wealth continued to favour lower odds of death.

### 3.5 Model diagnostics

Variance inflation factors for all covariates in the multivariable model ranged from 1.00 to 8.98, with the highest value observed for the no-education category of maternal education. No value exceeded the conventional threshold of 10, indicating that multicollinearity did not materially affect the stability of the adjusted odds ratios reported above.

## 4. Discussion

This nationally representative analysis of the 2022 Ghana Demographic and Health Survey found that children born less than 24 months after an older sibling, and especially those born within 18 months, faced substantially higher odds of death before age five than children born after a 24 to 35 month interval, even after adjustment for birth order, maternal age, plurality, residence, region, maternal education, household wealth, and marital status. The pattern observed here, in which risk was concentrated in the shortest intervals and levelled off beyond about three years, is consistent with the gradient described in the original multi-country Demographic and Health Survey analyses (2,3) and in the more recent international comparison (10), and it closely mirrors the magnitude of association reported for short intervals in the eight-country West African analysis that included Ghana (11) and in the Ghana-specific analysis of socioeconomic predictors of short spacing (13).

The mechanisms most often invoked to explain this association cannot be tested directly with the cross-sectional data used here, but each has a substantial empirical basis. Maternal depletion holds that a mother who conceives again before her physiological reserves have recovered from the preceding pregnancy and lactation enters the next pregnancy in a compromised state; the concept was formalised as the maternal depletion syndrome (27) and the nutritional burden it describes is considerable, with lactation itself raising energy requirements by roughly a quarter and protein requirements by more than half (28), and with the period of postpartum ovarian recovery shaped by maternal energy allocation (29). A short interval also curtails the duration of breastfeeding, since lactational amenorrhea is a major determinant of postpartum infertility and a subsequent conception displaces it (30). Alongside these biological pathways, the sibling competition theory holds that closely spaced children place competing demands on household resources, food, and caregiving time, and that the risk profile of a child born after a short interval may also reflect suboptimal breastfeeding of the preceding child or a maternal history of preterm birth (4). Direct transmission of infection between young siblings in the same household compounds these risks. That the association persisted after adjustment for socioeconomic and demographic confounders is in keeping with a genuine biological and behavioural pathway rather than simple confounding by social disadvantage.

The possibility of reverse causation nevertheless deserves explicit attention. A short interval can itself follow the death of a preceding child, when the resumption of fertility is accelerated by the loss of breastfeeding-related infecundability and, in some settings, by a deliberate wish to replace the child who died. Pooled data from 26 sub-Saharan African countries show the strength of this child replacement effect: the median subsequent birth interval was 24 months after a neonatal death and 25 months after a post-neonatal infant death, compared with 38 months after a child survived infancy, and prior neonatal death was associated with a 69% higher hazard of a subsequent birth and post-neonatal infant death with a 50% higher hazard (31). Comparable analyses in Bangladesh and rural Tanzania likewise find that the time to the next conception is shortened most when a child dies within the following interval, reflecting the combined operation of biological and volitional replacement, and that this response is stronger in high-mortality settings (32–34), with an estimated replacement effect of 0.37 in the Indian panel data (35). This well recognised mortality-replacement pathway means that some of the association estimated here may run from an earlier death to a short subsequent interval as much as from a short interval to a later death, and the data available cannot separate these two directions.

The strong and largely persistent regional variation is a further notable feature of the results. Even after adjustment for the individual and household characteristics available in this dataset, children in Volta, Northern, Western, Upper West, Ashanti, Savannah, and North East regions faced markedly higher adjusted odds of death than children in Greater Accra. This pattern is broadly consistent with previously documented geographic inequalities in Ghana’s under-five mortality (14) and with evidence on the geography of access to care. A national audit found that 34% of Ghanaian women of childbearing age live beyond the clinically significant two-hour threshold from facilities offering emergency obstetric and neonatal care at the partial standard or better, and 45% live that far from comprehensive facilities, rising to 63% and 81% respectively in the most remote regions (36). Access is markedly uneven between regions: 82% of women of childbearing age in Greater Accra can reach comprehensive emergency obstetric and neonatal care within two hours, compared with 19% in the mostly rural Northern region (37), and about half of Ghanaian women live two or more hours from facilities providing comprehensive obstetric care (38). This body of work suggests that factors not captured by the covariates used here, most plausibly the distribution of health facilities, the quality and accessibility of emergency obstetric and newborn care, and local patterns of water, sanitation, and nutrition, continue to shape a child’s chances of survival independently of birth spacing (39).

The elevated risk associated with multiple births is well established and is consistent with earlier pooled analyses of twin mortality across sub-Saharan Africa (40), which attribute much of this excess risk to the higher rates of preterm birth, low birthweight, and birth complications that accompany multiple pregnancies. Pooled data from 30 countries report roughly five-fold higher neonatal mortality in twins than in singletons, with about 60% of twins delivered prematurely, and estimate that around 10%, or 300,000, of the 3 million under-five deaths in sub-Saharan Africa occur in twins (41). A further synthesis of 156 surveys from 42 countries puts under-five mortality about three times higher among twins than singletons, with the excess concentrated in biomedical and nutritional features such as low birthweight, non-use of caesarean delivery, and lack of breastfeeding (42), and Eastern African meta-analytic evidence reports that 32% of twin pregnancies end in preterm delivery and 44% in low birthweight (43). The higher risk observed among births to mothers who had never been in a union may reflect reduced access to a partner’s material and caregiving support, although the confidence interval for this estimate was wide and the finding should be interpreted cautiously given the comparatively small number of births in this category.

Maternal age at the extremes of the reproductive range is frequently associated with child mortality elsewhere (44,45), and the crude excess risk observed here among births to mothers under 20 years of age is consistent with that literature, although it did not remain statistically significant after adjustment, most likely reflecting the moderate number of such births in this cohort combined with correlation between young maternal age and other covariates already in the model, such as birth order and marital status.

The absence of a statistically significant association between maternal education or household wealth and under-five death in the fully adjusted model should not be read as evidence that socioeconomic position is unimportant for child survival in Ghana. Both covariates showed a consistent gradient in the expected direction, and education reached significance in the unadjusted model; their attenuation after adjustment is most plausibly explained by their correlation with region and residence, which absorb much of their apparent effect once entered into the same model, and by the relatively small number of deaths available to estimate effects within the smallest wealth and education categories once other covariates are held constant.

## 5. Strengths and limitations

This study has several strengths. It used a nationally representative sample obtained through a rigorous, internationally standardised survey design, applied appropriate survey weighting and variance estimation throughout, and restricted the analytic cohort to births with a fully observed five-year survival window, avoiding the immortal-time bias that can arise when mortality outcomes are defined for children who have not yet completed the relevant age range. The multivariable model adjusted for a comprehensive set of demographics, socioeconomic, and geographic covariates identified in prior literature, and multicollinearity was formally assessed and found not to be a material concern.

Several limitations should also be acknowledged. First, the cross-sectional, retrospective design means that birth spacing and mortality were measured at a single point in time through maternal recall of past births and deaths, which can be subject to recall error, including possible heaping of reported ages at death or birth dates on round numbers; such error would be expected to attenuate rather than exaggerate the associations found. Second, as discussed above, the direction of causation between a short birth interval and child death cannot be established from these data, and mortality-induced replacement of a previously deceased child may contribute to part of the observed association (31). Third, biomedical and care-seeking covariates such as antenatal care attendance, place of delivery, and birthweight, which are recorded only for births in the five years immediately preceding the survey, could not be included because they are largely unavailable for the 5-to-10-year birth cohort used here; residual confounding by these factors cannot be ruled out. Fourth, the analysis was restricted to a binary under-five death outcome rather than a time-to-event framework, meaning that it could not account for the precise timing of death within the under-five period or for variation in the length of exposure to risk among children who died at different ages; a discrete-time or continuous-time survival approach was considered but not pursued in this analysis. Fifth, siblings born to the same mother are not statistically independent, and the design-based variance estimation accounts for clustering by enumeration area and stratum but not for the correlation of outcomes within a mother, so the reported confidence intervals may be somewhat narrower than a sibling-aware analysis would produce. Finally, as with all Demographic and Health Survey analyses, the exclusion of non-household populations and any underlying non-response mean that the sample, although nationally representative of the surveyed population, may not generalise perfectly to every subgroup of Ghanaian women and children.

## 6. Implications for practice and policy

These findings reinforce the continued relevance of the World Health Organization’s recommendation that women wait at least 24 months after a live birth before the next pregnancy (6), and they support sustained investment in postpartum family planning counselling and service provision within Ghana’s primary health care system, including through community-based health planning and services compounds, as a means of extending birth intervals and reducing under-five deaths (8,9). Evidence from low- and middle-income countries indicates that postpartum contraceptive use remains far from universal and varies widely across settings, so strengthening the supply of and counselling on postpartum methods is a necessary complement to demand-side messaging (46). Because unmet need for family planning in Ghana has itself been shown to vary geographically (47), family planning programming may benefit from being paired with the regional targeting suggested by the mortality patterns observed here, with particular attention to Volta, Northern, Western, Upper West, Ashanti, Savannah, and North East regions, where adjusted under-five mortality risk remained highest and where geographic access to emergency obstetric and neonatal care is most limited (36,37). The elevated risk identified among multiple births and among births to mothers who had never been in a union suggests that antenatal, obstetric, and postnatal services should maintain heightened clinical vigilance for these two groups specifically, including appropriate referral pathways for twin pregnancies and additional social support for unmarried mothers.

## 7. Recommendations for future research

Future research using Ghanaian data could usefully apply discrete-time or continuous-time survival models to the same birth-history data, which would allow the timing of death within the under-five period to be modelled directly and would make fuller use of information from children who have not yet completed a five-year observation window. Linking birth-history data to information on the distance to, and quality of, local health facilities would help to clarify how much of the regional variation identified here reflects health-system access rather than birth spacing or other individual-level factors, an approach that geospatial analyses of maternal and newborn care in sub-Saharan Africa have already shown to be feasible (39). Prospective cohort studies, which can establish the timing of birth intervals relative to subsequent child deaths with greater certainty than retrospective survey recall, would help to address the possibility of mortality-induced replacement raised above. Sibling-comparison designs, which contrast children of the same mother and so hold fixed the shared household and maternal characteristics that are difficult to measure, offer a further route to separating the effect of spacing from the confounding that retrospective designs cannot exclude (5). Finally, qualitative research into the barriers that Ghanaian women, and their partners, face in achieving a preferred birth interval would complement these quantitative findings and could inform more precisely targeted family planning programming.

## 8. Conclusion

In this nationally representative analysis of the 2022 Ghana Demographic and Health Survey, a short preceding birth interval, particularly one of less than 18 months, was independently associated with a substantially higher risk of under-five death, after accounting for birth order, maternal age, plurality, residence, region, maternal education, household wealth, and marital status. Mortality risk declined sharply as spacing lengthened from under 18 months to around 24 to 35 months, with no further evidence of a difference beyond that point. Multiple births and births to mothers who had never been in a union carried additional, independent risk, and substantial regional inequalities in under-five survival persisted after adjustment. Continued support for postpartum family planning, geographically targeted child survival programming, and closer clinical attention to multiple births and unmarried mothers are all indicated by these findings as means of reducing preventable under-five deaths in Ghana.

## 9. Abbreviations

AOR: Adjusted odds ratio
CI: Confidence interval
COR: Crude odds ratio
DHS: Demographic and Health Survey
GDHS: Ghana Demographic and Health Survey
GSS: Ghana Statistical Service
ICF: ICF International
OR: Odds ratio
PSU: Primary sampling unit
SDG: Sustainable Development Goal
STROBE: Strengthening the Reporting of Observational Studies in Epidemiology
U5MR: Under-five mortality rate
VIF: Variance inflation factor
WHO: World Health Organization

## 10. Declarations

### Ethics approval and consent to participate

The 2022 Ghana Demographic and Health Survey received ethical clearance from the Ghana Health Service Ethics Review Committee and the Institutional Review Board of ICF International, and written informed consent was obtained from all respondents at the time of the original data collection. The author accessed the fully de-identified Births Recode dataset for this registered secondary analysis under an authorisation granted by the DHS Program on 3 September 2026 (research project: “Birth spacing and under-five child survival in Ghana: evidence from the 2022 Demographic and Health Survey”).

## Consent for publication

Not applicable.

## Availability of data and materials

The Ghana Demographic and Health Survey 2022 Births Recode dataset analysed in this study is available from the DHS Program (https://dhsprogram.com) upon registration of a research project and approval of a data request. In keeping with the DHS Program’s terms of use, the author is not permitted to redistribute the micro-level data directly; researchers wishing to reproduce these analyses should request access to the Ghana 2022 Births Recode file directly from the DHS Program.

## Authors’ contributions

Abubakar Iddrisu Siddiq led the conceptualization, methodology, statistical analysis, and preparation of the original draft; Juliet Akpene Akuffo contributed to data curation and validation; Adu Gyamfi Oppong contributed to the literature review and interpretation of findings; Anne-Marie Xoese Ditchfield, contributed to data visualization and results interpretation; Sarah Agyarko Duah contributed to methodological review and critical revision of the manuscript; and Gideon Lantei Lamptey contributed to supervision, project administration, and final manuscript review, with all six authors approving the final version.

## Acknowledgements

The author thanks the DHS Program (ICF) and the Ghana Statistical Service for conducting the survey and making the data available for registered research use. Additionally, the author acknowledges the contributions of the Ghana Health Service in overseeing the ethical standards of the original survey implementation.

